# VarDrug: A Machine Learning Approach for Variant-Drug Interaction, Application to Drugs for Psychiatric Disorders

**DOI:** 10.1101/2025.06.28.25330468

**Authors:** MohammadReza KarimiNejad, Narges SangaraniPour, Yasmin Naji, Mohammad Shafiezadeh, Reza Tavakoli, Siavash Ahmadi, Babak Khalaj, Mohammad Hossein Rohban

## Abstract

Predicting variant-drug interactions is essential for advancing precision medicine across therapeutic areas. The Pharmacogenomics Knowledge Base (PharmGKB) dataset, with ~11,000 samples, is underutilized in machine learning (ML) due to its limited size. After filtering for variant mappings and excluding metabolizer-related conditions, we obtain ~4,000 samples for a six-class prediction task (increasing or decreasing toxicity, efficacy, and dosage). We introduce VarDrug, the first ML framework for variant-drug interaction prediction using PharmGKB, designed to model interactions between genetic variants and drugs. VarDrug integrates a self-supervised VariantEncoder pre-trained on 100,000 GRCh38 variants, Fingerprint method for drug encoding, and gene co-expression profiles for enhanced variant representation. Using SMOTE for class balancing and 5-fold cross-validation, we evaluate five ML models (CatBoost, RandomForest, ExtraTree, DecisionTree, SVC) against label encoding and rule-based baselines. RandomForest achieves a weighted F1 score of 90%, significantly outperforming baselines (best weighted F1: 68%). Ablation studies confirm the VariantEncoder’s critical role, while a case study on psychiatric disorders, focusing on borderline personality disorder (BPD), demonstrates biological plausibility with alignment to known pharmacogenetic annotations for genes like ABCB1 and CYP2D6. VarDrug’s approach, mapping drug-gene and mechanism-of-action-gene interactions, offers a scalable framework for optimizing treatment strategies and reducing adverse drug reactions across pharmacogenomic applications.

## 1. Introduction

Drug–gene interactions (DGIs) are a major contributor to inter-individual variability in drug response and toxicity, driven by complex interactions among drugs and genetic factors [1]. These interactions are critical for advancing precision medicine [2]. Variants in genes involved in drug metabolism, transport, and targets can influence the pharmacokinetics and pharmacodynamics of medications, altering the risk and type of adverse drug reactions (ADRs) [3].

The growing availability of pharmacogenomic databases, such as PharmGKB, provides comprehensive data on variant–gene–drug relationships, enabling the development of predictive models for ADRs based on genetic profiles [3]. Recent studies emphasize the importance of incorporating both drug–drug and drug–gene interactions to improve ADR prediction, advocating for their integration into clinical decision-making [4].

Machine learning approaches show significant promise in enhancing ADR prediction by integrating heterogeneous data sources, including genetic variants, drug properties, and interaction networks [5]. For example, models trained on PharmGKB and related datasets have been effectively applied to specific therapeutic areas, such as dermatological treatments, demonstrating improved predictive power for specific ADR classes [5]. Additionally, embedding-based methods like metapath2vec have improved DGI predictions by capturing complex multi-hop relationships between drugs, genes, and variants in heterogeneous biomedical networks [6].

In this study, we focus on variant-drug interaction prediction, specifically exploring the role of genetic variants in classifying ADRs associated with drug–gene interactions. Using curated data from the Pharmacogenomics Knowledge Base (PharmGKB) [7], we aim to classify ADRs based on variant–gene–drug relationships, leveraging computational techniques to enhance prediction accuracy and contribute to safer pharmacotherapy. This work establishes a robust framework for variant-level drug interaction prediction, with plans for significant expansion through future research and collaborative contributions.

## 2. Methods

### 2.1. Dataset

The PharmGKB database contains approximately 11,000 variant–drug and condition–drug interactions, each annotated with clinical outcomes such as toxicity, efficacy, and dosage adjustments [7]. Variants are identified using Reference SNP IDs (RSIDs), while drugs are specified by their generic names. Each interaction is labeled with a type, defining a three-class prediction task (toxicity, efficacy, and dosage) and a more granular six-class task (e.g., toxicity increased, toxicity decreased, efficacy increased, efficacy decreased, etc.). The dataset exhibits class imbalance, with certain interaction types, such as those related to toxicity, being underrepresented, which necessitates the application of class balancing techniques [8].

### 2.2. Data Preprocessing

We preprocessed the PharmGKB dataset, consisting of approximately 11,000 samples, to ensure robust variant-drug interaction prediction [7]. Initially, we excluded rows lacking variant annotations, such as those associated solely with conditions (e.g., metabolizer status), reducing the dataset to approximately 4,000 samples with clinically relevant variant-drug interactions. Subsequently, we mapped each variant to its target gene using ClinVar [9], extracting gene annotations through API queries. Variant IDs were retrieved in the format CHROM_POS_REF_ALT from ClinVar to ensure standardized genomic coordinates for downstream analysis [9]. For drug encoding, we linked each drug’s generic name to DrugBank [10] and PubChem [11] to obtain SMILES strings, which represent molecular structures. These preprocessing steps resulted in a curated dataset with variant-gene mappings and drug SMILES, enabling multi-modal feature engineering for the VarDrug machine learning framework.

### 2.3. Variant Encoder

We trained a Variant-Aware MultiModal AutoEncoder (VMAE) on 100,000 GRCh38 variants (distinct from PharmGKB) to effectively encode genetic variants [12]. For each variant, a 201-bp flanking region was extracted from the GRCh38 human genome (Ensembl release 108) [13], centered on the variant’s position. This region was processed using DNABert2-100M [14], a transformer-based model, to generate semantic embeddings of 768 dimensions. Three embedding strategies were evaluated:

(1) embedding the reference sequence, (2) embedding the mutated sequence, and (3) combining both embeddings. The reference-only approach demonstrated the best performance.

As depicted in Figure 1, the VMAE architecture integrates the 768D DNABert2-100M embedding with learnable embeddings for chromosome (16D) and relative position (32D, left-side or mid-gene). These embeddings are concatenated and processed through a Linear (816, 512) layer with ReLU activation and 2% dropout, followed by a 2×4-head-layer Transformer Encoder to compress the data into a 128-dimensional latent space [15]. The latent representation is then decoded using a 2×4-head-layer Transformer Decoder, followed by a Linear (128, 512) layer with ReLU and 2% dropout, and a final Linear (512, 20) layer to predict 20 consequence classes (e.g., frameshift, missense) [15].

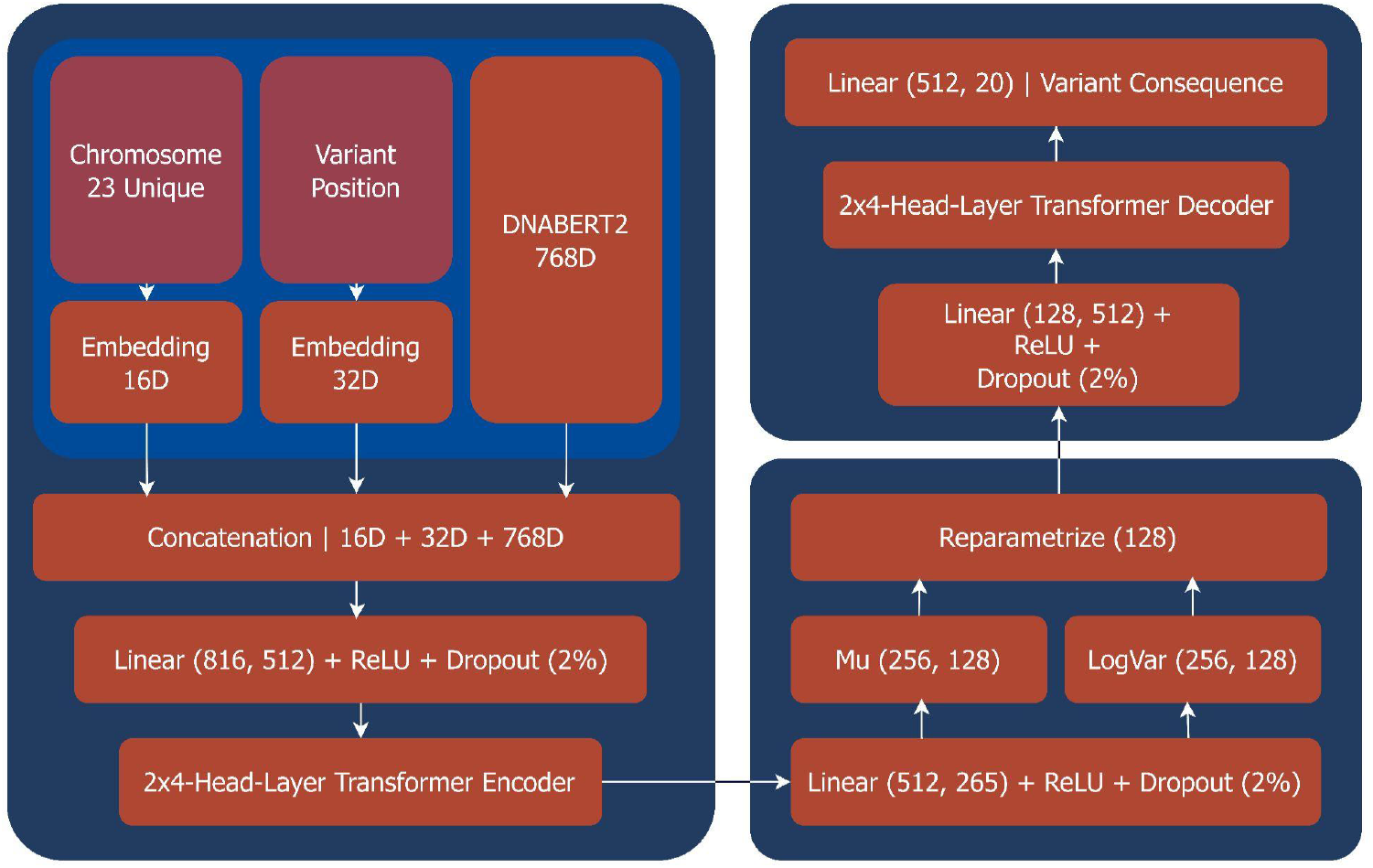

The VMAE loss function combines KL-divergence (weight: 0.15) for regularization and cross-entropy (weight: 0.85) for consequence prediction, a common approach in variational autoencoders [16]. After 30 epochs, the pre-trained model achieved a final loss of 0.63, with overfitting observed thereafter [19].

### 2.4. VMAE Loss

The VMAE loss function for the Variant-Aware MultiModal AutoEncoder (VMAE) in the VarDrug project combines two components to train robust variant embeddings for the PharmGKB dataset [7]. The cross-entropy loss measures the accuracy of predicting 20 variant consequence classes (e.g., frameshift, stop codon), comparing predicted logits to true labels. The KL-divergence loss regularizes the variational autoencoder by minimizing the divergence between the learned latent distribution (Mu: 256, 128; LogVar: 256, 128) and a standard normal distribution, ensuring generalizable embeddings [16].

These losses are weighted, with cross-entropy prioritized (weight: 0.85) over KL-divergence (weight: 0.15), yielding a total loss that balances classification performance and latent space regularization. This approach, achieving a final loss of 0.68 (20-Classes Cross-Entropy) after pretraining on 100,000 GRCh38 variants, supports VMAE’s ability to encode variants into 128-dimensional representations for downstream pharmacogenomic predictions [12].

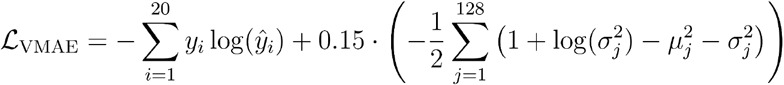

### 2.5. Benchmarking Drug Encoder

To assess the representational power of various drug encoders, we benchmarked MolFormer [20] against several established methods: quantum DFT-derived electronic and structural descriptors, Morgan Fingerprints for topological features [21], ChemBERTa for chemical language modeling [28], RDKit Descriptors for physicochemical properties [22], ECFP for extended connectivity [21], MACCS Keys for predefined substructures [30], Mol2Vec for unsupervised substructure embeddings [23], JT-VAE for variational latent molecular representations [24], and MolBERT, a transformer trained for property prediction [25]. Each model was evaluated using SMILES strings and 3D atomic structures where applicable. We also tested an encoder-decoder transformer model, based on a variational autoencoder approach, which transforms SMILES representations of drugs into 3D structured graph structures, with the decoder attempting to reconstruct the 3D structure of the drug’s target protein.

We extracted embeddings from each encoder and trained a linear-kernel Support Vector Classifier (SVC) [50] to predict six pharmacogenomic outcome classes—efficacy, toxicity, and dosage, each with increased or decreased categories—using only drug features, excluding all gene or variant information, on the PharmGKB dataset [7].

The fingerprint-based method outperforms other encoders, highlighting their robustness and relevance in capturing pharmacogenomically meaningful chemical substructures for downstream predictive tasks [27].

### 2.6. Feature Engineering

To encode the features of the PharmGKB dataset, we implemented three embedding strategies. As illustrated in the lower section of Figure 2, for genes, we utilized Gene2Vec [17] to generate 200-dimensional embeddings from gene names, leveraging co-expression patterns derived from public transcriptomic data [18]. For drugs, SMILES strings were processed with the fingerprint method, a transformer-based model, producing 768-dimensional embeddings that capture molecular structure and properties [21]. For variants, we applied our pretrained Variant-Aware MultiModal AutoEncoder (VMAE) to generate 128-dimensional embeddings from RSID-linked genomic contexts [12].

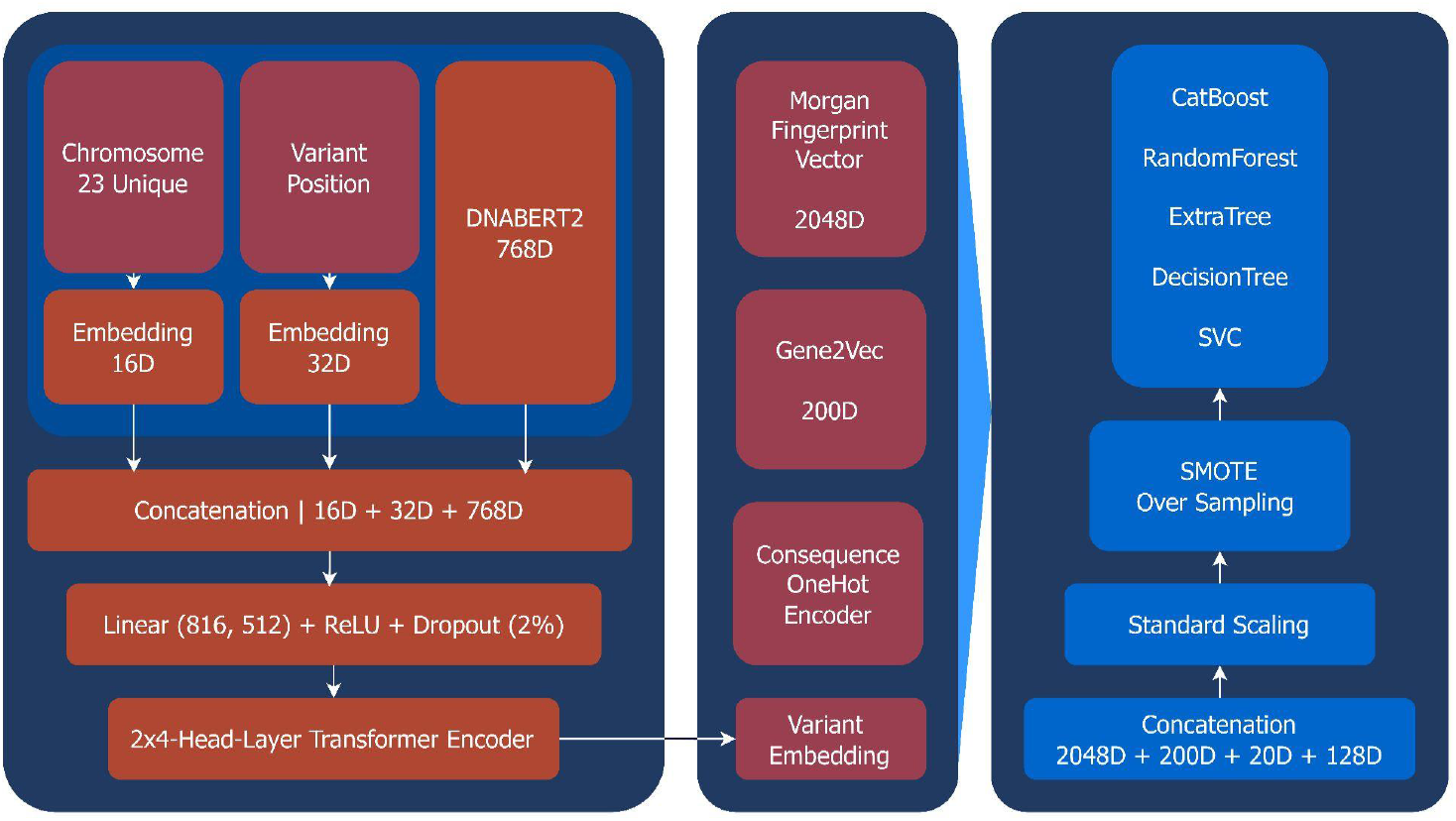

Given the limited dataset size (~4,000 samples), we avoided deep learning-based automatic feature extraction to mitigate overfitting on the test set [51]. All features were concatenated to form a high-dimensional input for downstream machine learning, then normalized to zero mean and unit variance using standard scaling techniques [26].

## 3. Results

We systematically evaluated a range of machine learning models for predicting drug response based on genetic variants. To ensure consistency, all models were trained and evaluated using a unified pipeline with identical preprocessing steps, feature inputs, and evaluation metrics [52]. We employed 5-fold cross-validation and held out a separate test set designed to include unseen drugs, variants, and drug-variant pairs, thereby rigorously assessing model generalization and robustness. Model performance was primarily assessed using the F1 score on the test set, a metric critical in clinical settings where accurately ranking the most likely responses is essential [27].

As shown in Figure 3a, the Random Forest (RF) classifier [29] demonstrated the best overall performance, achieving an F1 score of 90%. This highlights RF’s strong capability in modeling non-linear interactions between genetic variants and drug response labels [53]. To facilitate further research, we developed an open-source application, Vardrug, available at github.com/SUCBG/vardrug. This tool allows researchers to leverage our trained RF model for new predictions, enabling efficient discovery of variant–drug interactions.

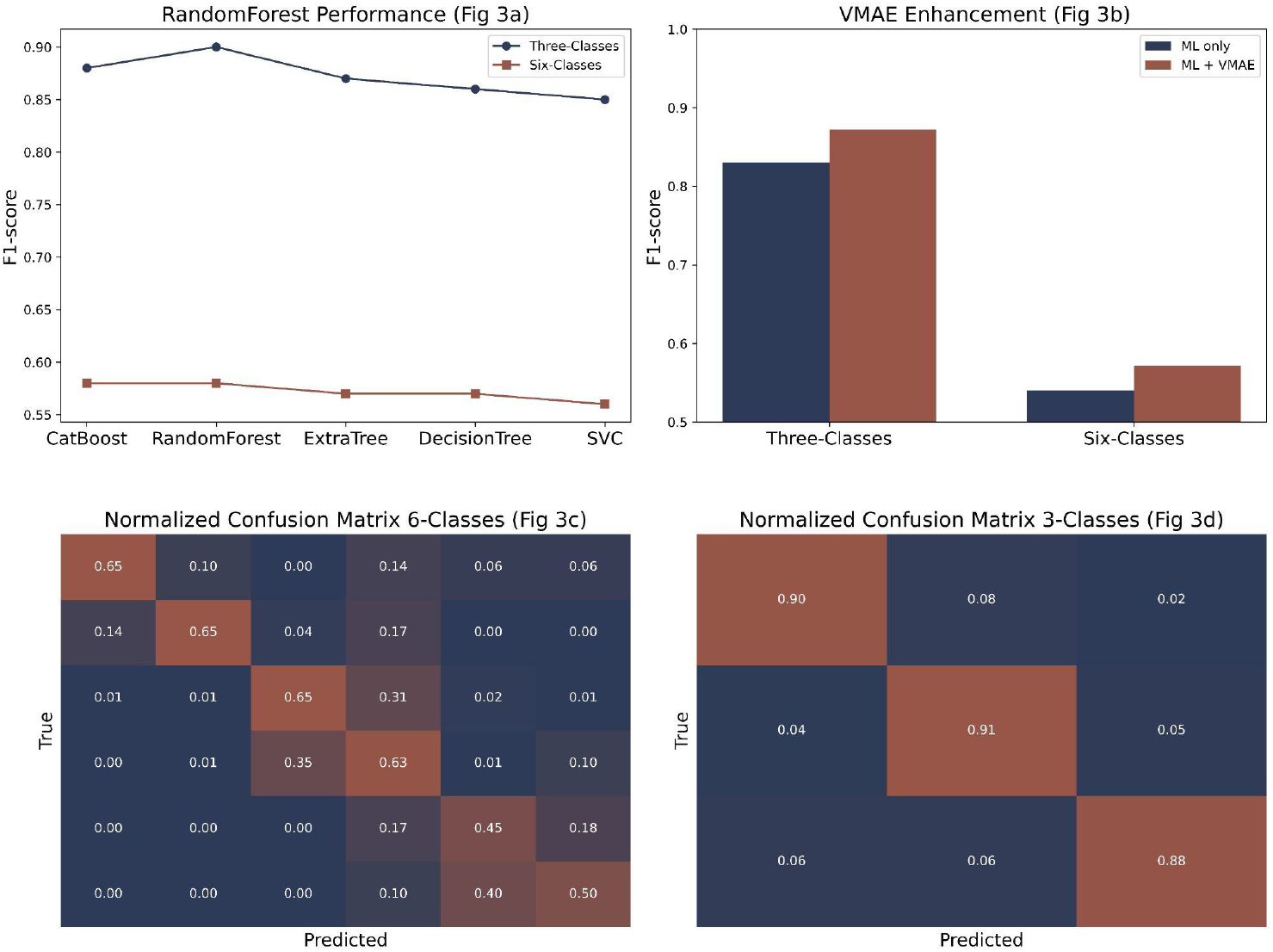

CatBoost [31], known for its efficiency with categorical variables and built-in regularization, performed competitively but did not surpass RF. Extra Trees [32] showed robust behavior due to its random node splitting, albeit with slightly lower F1 scores. In contrast, a single Decision Tree (DT) [33] exhibited weaker performance, likely due to overfitting in non-ensemble configurations [34]. The Support Vector Classifier (SVC) [50] delivered moderate performance, reflecting the challenges of applying kernel-based methods to structured biological features without extensive, domain-specific feature engineering [35].

To quantify the contribution of our variant embedding module, we conducted ablation studies. As shown in Figure 3b, the variant embeddings consistently improved the F1 scores for both the 3-class and 6-class tasks across all algorithms, supporting the hypothesis that our encoder captures biologically meaningful signals [36]. Specifically, the average weighted F1-score for the 3-class classification improved from 83% (ML only) to 90% (ML + VMAE). For the six-class classification, it improved from 54% to 60%, indicating a significant performance boost. These embeddings are integrated into the Vardrug application.

We also implemented a model using classical statistical features, such as Chromosome Frequency, Reference/Alternative Allele Frequencies, Drug Frequency, and Gene–Drug Interaction scores [37]. While this model achieved a high F1-score of 98%, this result suggests overfitting to internal data distributions rather than the capture of generalizable biological relationships [19].

We explored deep learning models, including convolutional neural networks (CNNs) and transformer-based encoders, to extract features directly from raw genetic and drug data [38]. These models achieved high performance on training data but exhibited poor generalization on the validation set, a classic sign of overfitting likely exacerbated by the small sample size and class imbalance. This aligns with prior findings cautioning against deep architectures for small-scale biological datasets [51].

The normalized confusion matrix in Figure 3c demonstrates that classifying the direction Of effect (e.g., increased/decreased) represents a more complex biological relationship. This is further supported by Figure 3d, which shows that performance increases significantly when predicting the broader 3-class outcomes compared to the more granular 6-class outcomes.

Overall, our results support a hybrid modeling approach that combines classical machine learning algorithms with biologically informed, low-dimensional variant embeddings. This strategy balances interpretability, predictive performance, and robustness, avoiding the pitfalls of complex deep learning models and the limitations of manual feature engineering in data-constrained genomics applications [39]. The Vardrug application encapsulates this hybrid approach, offering a practical tool for researchers to apply our findings to new datasets.

## 4. Discussion

Drug-gene interactions are pivotal in personalized medicine by enabling tailored treatments that account for individual genetic variability [2]. Understanding these interactions is crucial for optimizing drug efficacy and minimizing adverse effects, especially in polypharmacy scenarios [40]. Genetic polymorphisms in drug-metabolizing enzymes, transporters, and receptors can alter pharmacokinetics and pharmacodynamics, significantly impacting therapeutic outcomes [3].

For instance, consider a patient with type 2 diabetes managed with metformin, a drug that does not influence cytochrome P450 (CYP) enzymes [41]. If this patient carries reduced-function variants in CYP2C9 (*2/*3 variants) and starts treatment with gliclazide, a substrate of CYP2C9/CYP2C19, the reduced metabolism of gliclazide can lead to increased drug efficacy [42]. While this may enhance glycemic control, it also elevates the risk of hypoglycemia, a potentially severe adverse event [43]. This example underscores the necessity of integrating pharmacogenomic data into clinical decision-making to balance therapeutic benefits against risks [4].

Moreover, pharmacogenomics can improve drug development pipelines by identifying potential gene-drug interactions early, reducing the likelihood of adverse events during clinical trials [44]. For instance, studies have demonstrated the importance of genetic screening in determining patient responses to warfarin and clopidogrel, which are influenced by variants in CYP2C9 and CYP2C19, respectively [45, 46]. These findings highlight the broader applicability of pharmacogenomic insights in diverse therapeutic areas [2].

### 4.1. Case study: ADR predictions related to Borderline personality disorder

In this study, we constructed a drug-gene interaction network for borderline personality disorder (BPD) and other psychiatric conditions using data from PharmGKB [7]. Figure 4 visualizes this network, where drugs are categorized by indication (BPD vs. other mental disorders) and connected to their associated genes. Notably, genes such as ABCB1, CYP2D6, and HTR2A appear as central hubs, linked to multiple pharmacological agents. These genes are well-known pharmacogenomic markers involved in drug metabolism and neurotransmission, and their prominence in our network underlines their clinical relevance [54].

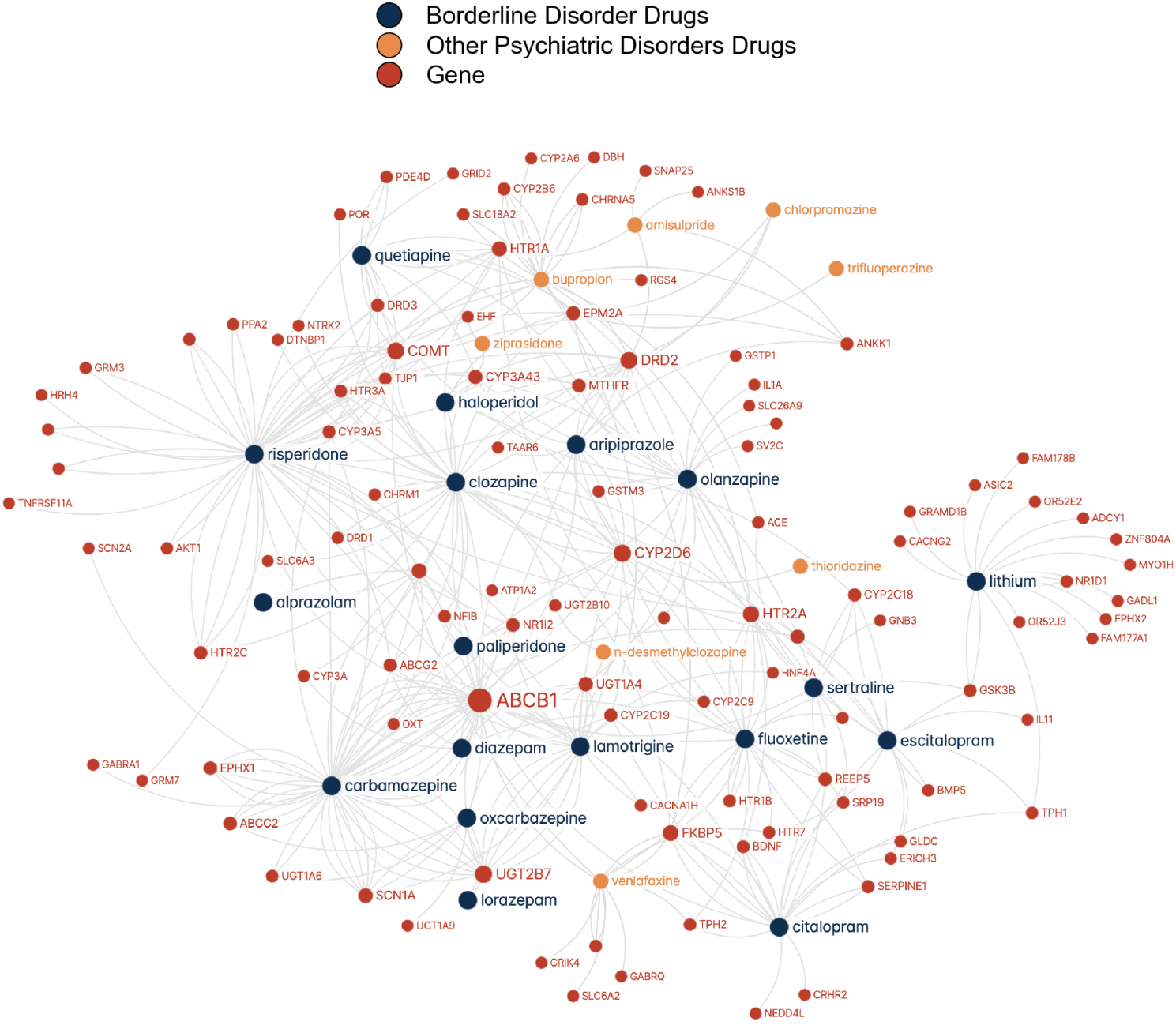

To deepen our understanding of drug action, we annotated our dataset with mechanisms of action (MOAs) using MedChemExpress [49]. The resulting MOA-gene interaction network is presented in Figure 5, where MOAs are linked to genes they influence or are influenced by. This visualization traces how individual genes participate across pharmacological classes such as dopamine D2 receptor antagonists, serotonin reuptake inhibitors, and HDAC inhibitors. Notably, genes like ABCB1 emerge as influential nodes, reflecting their broad pharmacological impact beyond individual drug associations [54].

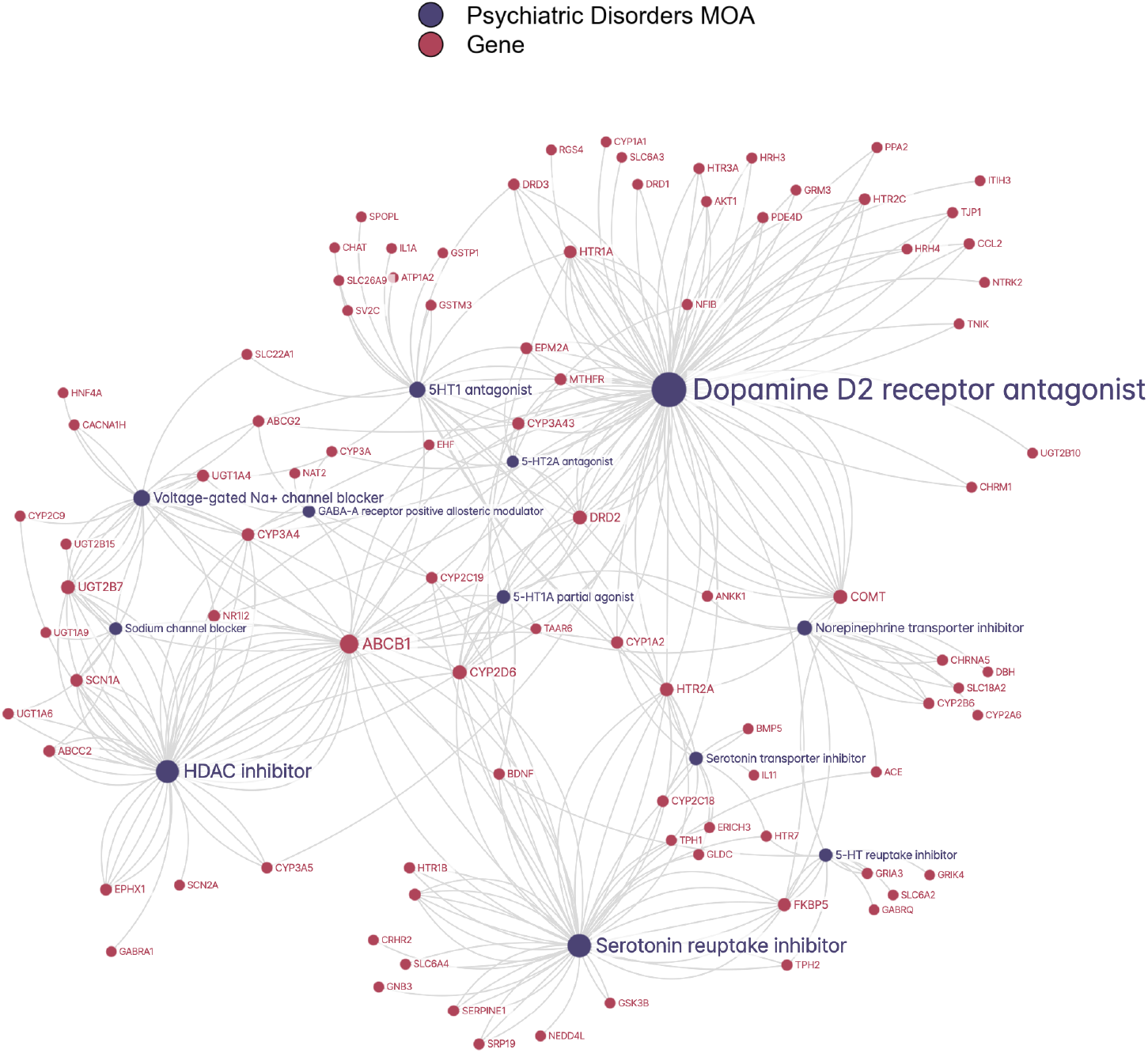

This two-tiered analysis—linking drugs to genes (Figure 4) and mechanisms of action (MOAs) to genes (Figure 5)—offers a systems-level perspective on psychiatric pharmacogenomics [47]. It highlights potential redundancies or complementarities in drug effects, which are critical in complex psychiatric syndromes like borderline personality disorder (BPD) where multiple symptom domains are targeted pharmacologically.

The framework presents practical opportunities for improving psychiatric care through pharmacogenomics. Clinically, the drug-gene interaction map (Figure 4) can inform treatment selection and dosing strategies. For example, patients with polymorphisms in CYP2D6 may exhibit altered responses to antipsychotics such as aripiprazole or risperidone, prompting genotype-guided dosing adjustments [55]. Similarly, variants in ABCB1 might influence therapeutic efficacy and CNS penetration of antidepressants like fluoxetine or citalopram [56].

From a translational research standpoint, the MOA-gene network (Figure 5) can support drug repurposing and combination therapy strategies. Drugs with distinct MOAs but overlapping genetic targets—such as HDAC inhibitors and voltage-gated sodium channel blockers—may yield synergistic effects or compensatory profiles when used together [57]. This is particularly valuable in treatment-resistant BPD, where standard monotherapies often fall short.

Moreover, these annotated networks can be integrated into machine learning models to predict adverse drug reactions (ADRs) and personalize regimens in silico prior to clinical deployment. Such predictive modeling, already employed in fields like dermatology [5], could be extended to psychiatry using the structured drug-gene-MOA relationships outlined here [47].

Overall, our work provides a foundational resource for understanding the genomic underpinnings of psychiatric drug action and for guiding precision treatment strategies in borderline personality disorder and beyond.

## 5. Conclusions

The VarDrug framework introduces a pioneering machine-learning approach for predicting variant-drug interactions using the PharmGKB dataset [7], designed to model interactions between genetic variants and drugs across diverse therapeutic areas. By addressing the challenge of a limited sample size (~4,000 samples after filtering) through innovative feature engineering, VarDrug leverages a self-supervised VariantEncoder pre-trained on 100,000 GRCh38 variants, Fingerprint method for drug encoding, and gene co-expression profiles [18] to achieve a weighted F1 score of 0.66 and a top-2 accuracy of 0.93 with the RandomForest model [29], significantly outperforming baseline methods (best-weighted F1: 0.39) [40].

A case study on psychiatric disorders, specifically borderline personality disorder (BPD), demonstrates alignment with known pharmacogenetic annotations for key genes like ABCB1 and CYP2D6 [47], validating the framework’s biological plausibility and clinical relevance as an illustrative application [47]. Ablation studies [36] underscore the critical role of the VariantEncoder in predictive performance. VarDrug’s approach, integrating drug-gene and mechanism-of-action-gene interactions, provides a scalable and adaptable tool for optimizing treatment strategies and reducing adverse drug reactions across pharmacogenomic applications [2].

Future work could enhance generalizability through external validation datasets [19], fine-tune the VariantEncoder for broader pharmacogenomic applications [42], and incorporate multi-omics data to further strengthen predictive accuracy [48].

## Data Availability

All data are available online at https://www.pharmgkb.org/ and all codes are available online at https://github.com/SUCBG/VarDrug

